# Bringing public attention to disease into global health priority-setting

**DOI:** 10.64898/2026.08.01.26359468

**Authors:** Wenceslao Arroyo-Machado, Ismael Rafols, Adrian A. Diaz-Faes

## Abstract

**Background:** A central concern in global health priority-setting is whether the supply of scientific knowledge aligns with health needs and demands. This alignment is usually assessed by comparing research effort with disease burden, overlooking other type of “social demand” of disease, in particular whether diseases are socially visible and generate public attention. We develop an analytical framework that treats public attention and epidemiological burden as complementary dimensions of health demand and examines their alignment with knowledge supply.

**Methods:** We combine data on publications indexed in OpenAlex, disability-adjusted life years from the Global Burden of Disease, and Wikipedia pageviews for 2016 to 2023, as indicators of research effort, disease burden, and public attention, respectively. We map 19 disease groups and 138 specific diseases across these three dimensions. Ternary plots are used to position diseases according to their relative balance across dimensions and to identify diseases that are over- or under-represented in research effort relative to epidemiological burden and public attention. We compare Global North-South patterns using German, Persian, Swahili, and Vietnamese language areas to assess how these relationships vary across territories.

**Results:** The three dimensions show limited alignment. At the disease group level, cardiovascular diseases account for the largest share of disease burden, mental disorders attract the largest share of public attention, and neoplasms concentrate the largest share of research effort. Public attention and disease burden are weakly correlated at both group and specific disease levels, indicating that Wikipedia pageviews and DALYs capture distinct dimensions of health demand. Ternary plots reveal different forms of misalignment, with some diseases showing plots dominated by burden, others by research effort, and others by public attention. Territorial analyses add a further layer by showing that diseases follow disparate patterns of supply-demand (mis)alignment across different linguistic territories.

**Conclusions:** Public attention provides a complementary dimension for mapping global health needs and demands. Our approach identifies where scientific knowledge supply fails to match epidemiological and/or public attention, supporting more nuanced global health analysis that may be useful for priority-setting.

## 1. Background

Global health policy increasingly recognizes that health systems should be organized around the comprehensive needs of people and communities [1]. This principle for health priority-setting means that diseases impact not only in terms of the epidemiological burden they produce, but also in how they disrupt everyday life, become visible, and generate concern or information-seeking behaviours. Yet this social dimension is rarely incorporated into frameworks used to address health needs.

The Global Burden of Disease (GBD) framework and the disability-adjusted life years (DALYs) indicator have been central to the analyses of global health needs because they allow morbidity and mortality to be compared across diseases, countries, and regions [2–5]. This comparability has made visible important mismatches between health loss and research efforts. Existing evidence shows that the “supply” of knowledge through medical research, pharmaceutical innovation, and funding does not align with societal and health “needs and demands” in terms of disease burden. Instead, a variety of studies has shown that knowledge production is shaped by geographical factors, research capacity, market incentives, and institutional agendas that do not align well with health needs [6–14].

The objective of this study is to develop an analytical framework that investigates the alignment between science and health demands, using as indicators of health demands not only disease burden, but also public attention to disease to identify areas of potential need in global health. To do so, we first map diseases from the GBD classification across three dimensions, describing how research effort (knowledge supply), disease burden, and public attention are distributed across diseases.

We use the number of scientific publications as a proxy for research efforts, the number of DALYs as a measure of disease burden, and Wikipedia pageviews as an indicator of public attention to disease. Second, we examine the relationships between the three dimensions across broad disease groups and then at the level of specific diseases. These two steps provide the basis for the methodological contribution of the paper: the use of ternary plots as a mapping technique that allows the comparison of knowledge supply on one axis against knowledge demands on the other two axes (burden and public attention), allowing the visualisation of three dimensions to be compared within a common analytical space. This reveals different types of (mis)alignment, as diseases appear over- or under-represented in different dimensions.

We propose that public attention to disease provides a complementary dimension for mapping global health needs and demands. The GBD^1^ captures epidemiological burden (loss of life or disability) but does not show which diseases generate concern, uncertainty, stigma, or information-seeking among the public [15,16]. Incorporating public attention can therefore help capture aspects of health demand that remains overlooked when the analysis of priority-setting focuses only on the alignment between burden and research effort.

The paper is organized as follows. Section 1 develops the conceptual background by discussing disease burden and global health prioritization, the social aspects of disease, and the use of Wikipedia as a source for mapping public attention. Section 2 describes the data, indicators, and the ternary plot design used to compare the three dimensions. Section 3 presents the results, and Section 4 discusses the implications of the framework for identifying priority areas in global health policy.

### 1.1. Prioritization of global health efforts

A persistent challenge in global health policy is deciding how limited resources should be distributed across diverse health needs. Health systems, public agencies, and research funders have often addressed this challenge through disease-specific, or vertical, approaches, organizing priorities and interventions around particular diseases or disease groups associated with greater health loss [17,18]. This perspective follows the logic proposed by Sarewitz and Pielke [13] that science “supply” should be aligned with societal (in this case health) “needs and demands”.

The development of the GBD framework and DALYs indicator has been central not only to estimate disease burden but to help prioritize efforts on disease on a global scale [2,3]. By combining mortality and morbidity into a single indicator, DALYs makes disease burden comparable across countries, regions, and disease groups [4,5]. For instance, Coburn et al. [19] argue that neglected diseases gained policy momentum because DALYs made the misalignment between science supply and demand visible: some diseases caused a lot of health loss but received very little research funding. This helped turn neglected diseases into a clear case for targeted R&D investment [20,21]. Despite major efforts, a body of empirical work shows that disease burden and research effort remain strongly misaligned and shaped by intertwined governance, economic, and social factors [9,22–25].

Evans et al. [7] show that global research efforts do not follow DALYs but mainly respond to some diseases with high burden in Global North countries. Since high-income countries produce a large share of biomedical research, diseases prevalent in these countries tend to receive disproportionately more scientific and clinical research efforts. Yegros-Yegros et al. [9] extend these findings, reporting that diseases with comparable epidemiological burden in high- and middle-income countries are also significantly under-researched. Kumar et al. [26] show that this misalignment also appears within countries. In India, cardiovascular and respiratory diseases account for a large share of disease burden but receive relatively little research efforts, while cancer receives a much larger share of publications than would be expected from its burden. Schmallenbach et al. [10] add that recent improvements in burden-research alignment may not be due to better policies, as the increased alignment seems to be driven mostly by the decline of communicable diseases and the spread of non-communicable diseases, rather than by a shift in research priorities. This misalignment also extends to how clinical evidence is produced [27]. Lou et al. [11] show that participation in randomized control trials is shaped far more by country-level research capacity, health infrastructure, and governance than by disease burden.

These misalignments are reinforced by the way global health research and innovation are funded. A large share of health research and innovation funding comes from industry: around 60% of health R&D investment in high-income countries comes from the business sector [23]. This matters because pharmaceutical innovation is strongly influenced by market demand and expected return-on-investment, with research concentrated in diseases that offer larger commercial opportunities [9,28]. The pattern is different for neglected and poverty-related diseases [24], where research and innovation investment depend mainly on public, multilateral organizations (i.e. WHO, World Bank, The Global Fund) and philanthropic donors [29,30]. Philanthropic funding plays a significant role in areas with weaker commercial incentives but it also brings its own agenda [31]. Charani et al. [25] show that large donors often channel grants through institutions in high-income countries. For instance, around 88% of funds of Bill & Melinda Gates Foundation are granted to Global North institutions, at the expense of R&D capacity building in the Global South. This can limit epistemic diversity in the definition of research agendas and reinforce particular ways of addressing health needs, as argued by Birn [32] who points out that philanthropy often favours industry and technical approaches to global health.

Taken together, this evidence shows that, despite funding and policy efforts to improve global health, research and innovation still often do not match the distribution of disease burden. This misalignment reflects how global health priorities are shaped by governance structures, markets, and institutional agendas [9,33]. Yet prioritizing health needs only through disease burden and funding constraints leaves out some of the social dimensions of disease, including the extent to which diseases become objects of public attention and information-seeking, which can be understood as a type of health needs or demands. This matters for priority-setting because public attention may reveal aspects of disease salience (of health demands) that are not captured by DALYs alone. The challenge is therefore not merely to document misalignments, but to develop methods that help identify, in a more comprehensive way, which dimension(s) of misalignment can be observed so as to inform further prioritization.

### 1.2. Approaching disease from a social lens

This broader view of the social dimension of disease should matter for health prioritization because diseases impact not only in the health losses they produce, but also in how they disrupt everyday life, shape lived experience, and become recognized as matters of public concern. A disease may impose substantial health losses while remaining weakly visible and accepted in public debate, or it may generate public concern, stigma, or information-seeking beyond what its epidemiological burden alone would suggest [16,34]. For this reason, public attention to disease can complement epidemiological burden to indicate health needs and demands.

These social aspects are closely related to the difference between disease and illness. Disease refers to the pathological condition diagnosed and treated by medicine, whereas illness refers to the lived experience of discomfort, dysfunction, and changes in social functioning by an individual [15]. They can affect identity, work, relationships and everyday life. Conditions such as mental disorders, skin diseases, breast cancer, chronic pain, or some neglected tropical diseases may therefore carry forms of stigma, fear, visibility, or disruption that are not well captured by DALYs [16,34–36]. Neglected tropical diseases such as lymphatic filariasis, leprosy, or leishmaniasis illustrate this point: their impact is reflected not only in years of life lost, but also in long-term and often visible effects, including disfigurement, disability, stigma, and social exclusion, with consequences for everyday life and access to care [20,37]. For instance, health-related stigma has been described as a hidden burden of illness: a social process through which individuals or groups identified with particular health problems are rejected or devalued in ways that may affect help-seeking, treatment, and policy response [38]. Evidence from mental health shows that greater acceptance of neurobiological explanations can increase support for treatment without reducing stigma [39].

### 1.3. Mapping public attention to disease through Wikipedia

One way in which the social dimension and the “demand” about a knowledge of disease become visible is through information-seeking behaviour. When diseases generate interest, stigma, fear, media coverage or public debate, people may seek information about them to make sense of symptoms, diagnoses and risks. Online information-seeking can therefore be understood as an expression of public attention to disease: it captures the extent to which a condition becomes salient enough for people to search, read and learn about it.

Public attention to disease can provide a useful complement to DALYs because it captures aspects of health needs and demands and of disease salience that are not directly measured through GBD. This is increasingly relevant as access to information and communication technologies expands globally. Recent estimates shows that internet use in Africa has reached around 40% of the population and is expanding largely through mobile broadband [40,41]. Thus, public attention to disease can help identify social dimensions of disease that may otherwise remain weakly represented in priority-setting frameworks.

In this study, we use Wikipedia to proxy for public attention to disease, which we propose to relate to the ‘demand’ of knowledge following Pielke and Sarewitz (2007). Wikipedia is a free, collaborative online encyclopaedia and consistently ranks among the most visited websites worldwide. It is one of the most visible and frequently used online health information resources for the public, patients, students and practitioners, with health-related articles often ranking highly in search engine results [42,43]. Although access is totally o partially restricted in some countries, such as mainland China and Saudi Arabia^2^, Wikipedia has proven valuable for public health research and health monitoring, including studies of HIV, malaria and influenza [44–47]. As of June 2026, 345 Wikipedia language editions were active. Qualitative evidence also suggests that users turn to Wikipedia because it is familiar and convenient, and because it serves as a starting point for understanding health issues [42]. Regarding the quality of its content, studies point out that Wikipedia’s accuracy is comparable to that of conventional encyclopaedias and that article quality depends not only on contributor types but also on patterns of collaboration [48].

Public attention to disease, measured through Wikipedia, can be studied together with epidemiological burden and research efforts through novel mapping and visualisation techniques. Mapping techniques are useful for health prioritization because they make visible how different dimensions of a problem relate to one another. In science and innovation studies, maps have been used to reveal the dynamics of biomedical innovation [49,50], highlight knowledge and funding structures [51,52], or detect research priorities against societal needs [12,53]. In this paper, we apply this approach to global health prioritization by using ternary plots to position diseases according to their relative balance across epidemiological burden, research effort, and public attention. Note that maps should not be read as prescriptive, rather they make visible distributions across three dimensions, which show patterns of (mis)alignment that can guide further reflection and context-specific decision-making.

## 2. Methodology

### 2.1. Database construction and indicators

Our analytical framework integrates data from three dimensions: research effort, epidemiological burden, and public attention. Each dimension is captured through distinct data sources and processed separately. Data for all three dimensions span the period from 2016 to 2023, based on the availability of Wikipedia pageview statistics (from mid-2015 onwards) and the latest GBD data (2023). We conducted the manual matching of GBD diseases to MeSH descriptors and Wikipedia articles in Microsoft Excel 365. Subsequent data processing and statistical analyses were performed in Python 3.13.5 and data visualization in R 4.5.1 through RStudio.

#### Science supply: Research effort through OpenAlex

Research effort is operationalized through the volume of scholarly publications addressing each disease. MeSH terms were manually assigned to each GBD cause, and subsequently cross-referenced with Yegros-Yegros et al. [9], which links GBD causes to one or more MeSH term. This comparative process enabled validation of the initial manual assignments and identification of additional relevant terms. Then, publications were retrieved from OpenAlex, limiting the sample to articles, reviews, and letters (i.e. citable items) in which the corresponding MeSH terms appear as a major descriptor. This way we ensured that only publications addressing a disease as a primary focus are included.

This process retrieved 1,470,273 unique publications. Note that when aggregating publications across hierarchical levels, we merged parent and child GBD categories and avoided duplication by using OpenAlex. For example, publications indexed under the level-3 disease *leishmaniasis* are aggregated with those under its level-4 subcategories (*visceral leishmaniasis* and *cutaneous and mucocutaneous leishmaniasis*), but only unique publications are counted for *leishmaniasis* as a whole. We followed the same procedure when aggregating to level-2 diseases, ensuring that publications addressing multiple related diseases are counted only once at each level of analysis.

#### Health demands: Epidemiological burden through the Global Burden of Disease

We start building our dataset from the GBD 2023 open database, published in 2025 by the Institute for Health Metrics and Evaluation [54]. The GBD comprises 375 diseases, of which 333 form the basis of this study after excluding injury-related causes. These are organized into a four-level hierarchical structure, enabling analysis at multiple levels of granularity. Level-2 comprises 19 diseases belonging to the two major level-1 categories: *communicable, maternal, neonatal, and nutritional diseases* (7 diseases) and *non-communicable diseases* (12 diseases). As these level-2 diseases represent broad categories (e.g., *cardiovascular diseases*, *mental disorders*), our design also targets the more specific diseases at levels 3 (143 diseases) and 4 (133 diseases). Table 1 summarizes the filtering applied to the GBD 2023 dataset to select such specific diseases and their matching to MeSH terms and Wikipedia articles. We conduct analyses at both level-2 and level-3, aggregating lower levels where applicable, to provide a more comprehensive basis for global health prioritization. This aggregation was applied to broad level-3 diseases (e.g. *intestinal nematode infections*), which could not be directly matched in Wikipedia, but could be reconstructed by aggregating their level-4 diseases (e.g. *ascariasis*, *trichuriasis*, and *hookworm disease*).

**Table 1.** Summary of GBD 2023 data filtering and matching to MeSH and Wikipedia.

| **GBD hierarchical level** | **Total**<br>All GBD causes | **Diseases**<br>Excluding injury-related causes | **Specific diseases**<br>Excluding broad levels and residual* diseases | **Matched diseases**<br>Specific diseases matched to MeSH/Wikipedia |
| --- | --- | --- | --- | --- |
| Level-1 | 3 | 2 | --- | --- |
| Level-2 | 22 | 19 | --- | --- |
| Level-3 | 176 | 158 | 143 | 129 (90%) |
| Level-4 | 174 | 154 | 133 | 107** (80%) |
| **Total** | **375** | **333** | **276** | **236 (86%)** |

\*Residual disease groupings such as *other cardiovascular and circulatory diseases*.

Epidemiological burden is quantified using disability-adjusted life years (DALYs) retrieved from the GBD 2023. DALYs provide a standardized measure of overall disease burden, expressed as the number of years lost due to ill-health, disability, or early death. DALYs at level-2 represent the sum of their corresponding level-3 diseases, which in turn aggregate level-4 diseases. Although not free of limitations [2], DALYs are the primary indicator used to proxy for disease burden. They have been extensively discussed in the literature and refined through regular revisions and expert consultation [3].

#### Health demand: Public attention through Wikipedia

Public attention to disease is measured through Wikipedia pageviews. We started by matching each disease with its corresponding primary English Wikipedia article. Where a disease was not represented in English Wikipedia, other linguistic editions were reviewed to provide a more comprehensive global view of public attention to disease. The identification process was done by cross-referencing MeSH terms, as many diseases have their corresponding MeSH terms embedded in Wikipedia articles or in the associated Wikidata entries. We included only articles that focused directly on the disease, rather than articles where the disease was mentioned only as part of a broader topic. For instance, for *malaria*, the main article is the one named after the disease itself^3^, whereas related but secondary articles (e.g., *malaria vaccine*) are excluded. Also, Wikipedia articles that mention a disease only in passing or address it within a broader context without making it the primary focus were excluded. This logic parallels the publication retrieval criterion, where only major MeSH terms are considered.

Through this process, we retrieved pageviews from all Wikipedia articles for all linguistic editions of each matched article via the Wikimedia REST API. As with publications, pageviews are aggregated following the GBD hierarchy, by summing views across all linguistic editions for level-3 and level-4 articles when analysing level-2 diseases, and across level-4 articles when analysing level-3 diseases. We have matched 236 specific diseases, including 129 level-3 and 101 level-4 diseases matched across all three dimensions, plus four additional level-4 diseases matched only to MeSH terms and another two matched only to Wikipedia articles. Note that our analysis was conducted at two levels. First, level-2 (disease groups) is the main level and is built by aggregating the level-3 and level-4 categories below it, covering 19 major disease categories. Then, level-3 (specific diseases) combines the diseases at this level with those inherited from level-4, covering 138 disease categories.

Since Wikipedia pageview data are available by language edition rather than by country, we investigate territorial dynamics using four language editions (German, Vietnamese, Persian, and Swahili) chosen to capture variation across world regions and Global North-South differences. For each case, DALYs data were filtered by the countries where the language holds official or national status, publications were restricted to those with first author affiliated with an institution in the selected countries [10], and Wikipedia pageviews were counted only from the matching language edition. Countries where the selected languages have official status are Germany (German), Vietnam (Vietnamese), Iran, Afghanistan, and Tajikistan (Persian), and Tanzania, Rwanda, the Democratic Republic of the Congo, Kenya, and Uganda (Swahili).

#### Reliability analysis

The reliability of the procedure was assessed through comparisons with datasets built in previous work. For publications, we validated our disease assignments against the dataset by Schmallenbach et al. [10]. They linked 8.6 million disease-specific publications to GBD level-2 categories using a triangulated approach (large language model classification, matching International Classification of Diseases codes with MeSH terms, and manual validation by medical experts). Among the publications present in both datasets, at least one of our level-2 assignments matched theirs in 94% of cases. Their classification, however, excludes the level-2 diseases *mental disorders*, *other infectious diseases*, and *other non-communicable diseases*, so that publications in these categories could not be validated. Besides, where Schmallenbach et al. [10] assigned multiple categories to the same publication, we did not treat additional categories in their classification as mismatches. An assignation was considered valid if our assigned category or categories were included among theirs, without requiring a complete match. Despite the methodological differences, the near-complete overlap observed between both approaches supports the reliability of our research design. For Wikipedia, no comparable external matching exists, as this is the first study to systematically link the full GBD classification to Wikipedia disease articles.

### 2.2. Statistical analysis and ternary plots

We first examine the distribution of, and associations, between the three dimensions. We run descriptive analyses at GBD levels-2 and-3, followed by a pairwise correlations analysis using Kendall’s rank correlation coefficient (τ). Kendall’s τ was selected over Pearson or Spearman because it is more robust to outliers, non-normal distributions, and performs well with small sample sizes (level-2: n = 19). We computed correlations across two levels to ensure consistent findings: (1) across the 19 level-2 categories, and (2) across 138 level-3 diseases with complete data. Values close to 1 indicate strong agreement in the ranking of diseases, whereas values close to 0 indicate substantial disagreement across them.

Then, we employ ternary plots to map each disease according to the relative distribution of its values across the three dimensions [55]. A ternary plot represents each disease as a point within an equilateral triangle whose vertices correspond to the epidemiological burden (DALYs), public attention (pageviews), and research effort (publications) dimensions, with the position of each point determined by the proportional share of each metric relative to all other diseases. (Fig. 1). By placing research efforts in the top vertex, the vertical dimension of the ternary plot represents balance between research supply (top) and health demands (bottom). The horizontal axis represents the type of health demand observed: a position towards the right signals relatively more disease burden than public attention, whereas a position towards the left signals diseases with more public attention than burden.

**Fig. 1.**
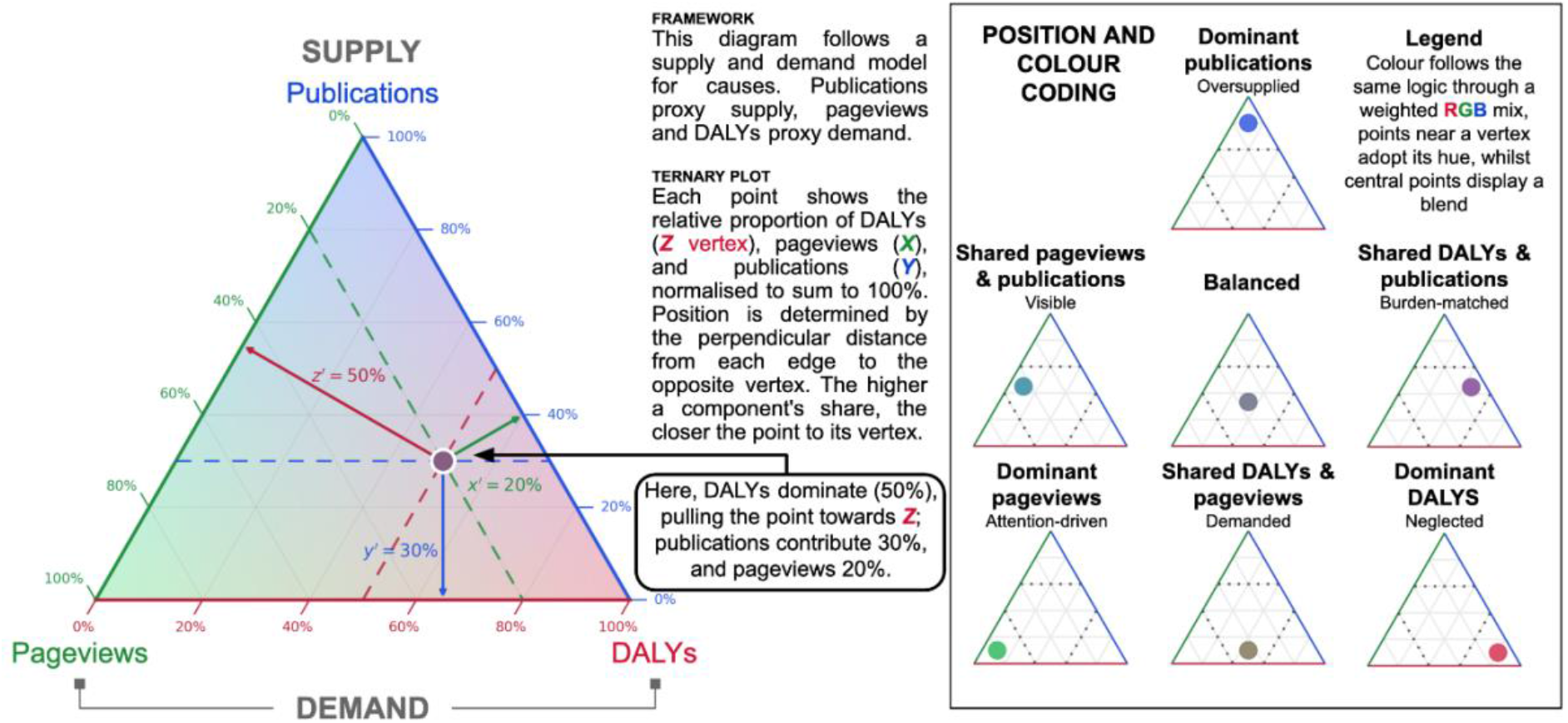
Reading guide for the ternary plots, illustrating how each disease location reflects the relative distribution of DALYs, pageviews, and publications, and the resulting profiles

We normalized the raw values before constructing the ternary plot because the three indicators are measured in different units. Each GBD disease’s value is divided by the total sum of that variable across all diseases. This normalization is preferred over min-max scaling, which forces at least one observation to 0 and another to 1, distorting the relative positioning of diseases as with small but meaningful values. Formally, for a given disease *c* and dimension *d* ∈ {DALYs, pageviews, publications}, the position of *c* in the ternary plot is defined as:

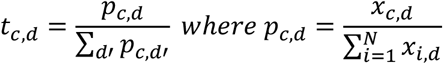

where *x_c,d_* denotes the raw value of disease *c* in dimension *d* and *N* is the total number of diseases. In other words: p_c,DALYs_ is the proportion of DALYs of disease c, p_c,Pageviews_ and p_c,Pubclications_ are proportions of pageviews and publications of the same disease. We then conduct a second normalisation, on the relative weight of a dimension (DALYs, pageviews, publications) in comparison to the other dimensions, which shows if a certain dimension is high in comparison to others.

For example, since *Neoplasms* make up 10% of all DALYs, 10% of pageviews and 31% of publications, in the ternary plot, they will be presented as 20% (10/51) for DALYs, 20% (10/51) for pageviews and 60% (31/51) for publications. Put differently, the ternary plots shows the relative weight of a disease in comparison to other dimensions: a disease near a vertex place most of its weight on one dimension (dominant), a disease near an edge between two vertices weight is shared between the corresponding two dimensions (shared), and a disease near the centroid weight is evenly distributed across epidemiological, research effort and public attention (balanced) (Fig 1).

Ternary plots were constructed for level-2 and level-3 diseases and for each level-2 disease group. To capture both the overall and within-group patterns, we present ternary plots in two modes. In the *absolute* mode, proportions are calculated against all diseases in the dataset, preserving each level-3 disease’s position relative to the full landscape of diseases. In the *relative* mode, proportions are recalculated within each disease group (level-3), revealing the relative importance of each disease within that specific group of diseases.

## 3. Results

### 3.1. Distribution, magnitudes, and differences across diseases

Fig. 2 provides an overview into how DALYs, scientific activity, and Wikipedia pages views are distributed across the 19 major disease areas (level 2). Global epidemiological burden is concentrated mainly in *cardiovascular diseases* (17% of total DALYs), *neoplasms* (10%), and *respiratory infections and tuberculosis* (9%), which together account for over one-third of global disease burden. Public attention distributes differently, with *mental disorders* in first position (11% of total pageviews), followed by *neoplasms* (10%). Research effort shows a more skewed profile, with *neoplasms* accounting for 31% of total Open Alex publications (even higher than in previous estimates of 25% using Web of Science [10,26]) followed by *respiratory infections and tuberculosis* (12%) and *cardiovascular diseases* (9%). Several disease areas show alignment across the three dimensions, as is the case for neurological disorders (4-6%) and *respiratory infections and tuberculosis* (9-12%).

**Fig. 2.**
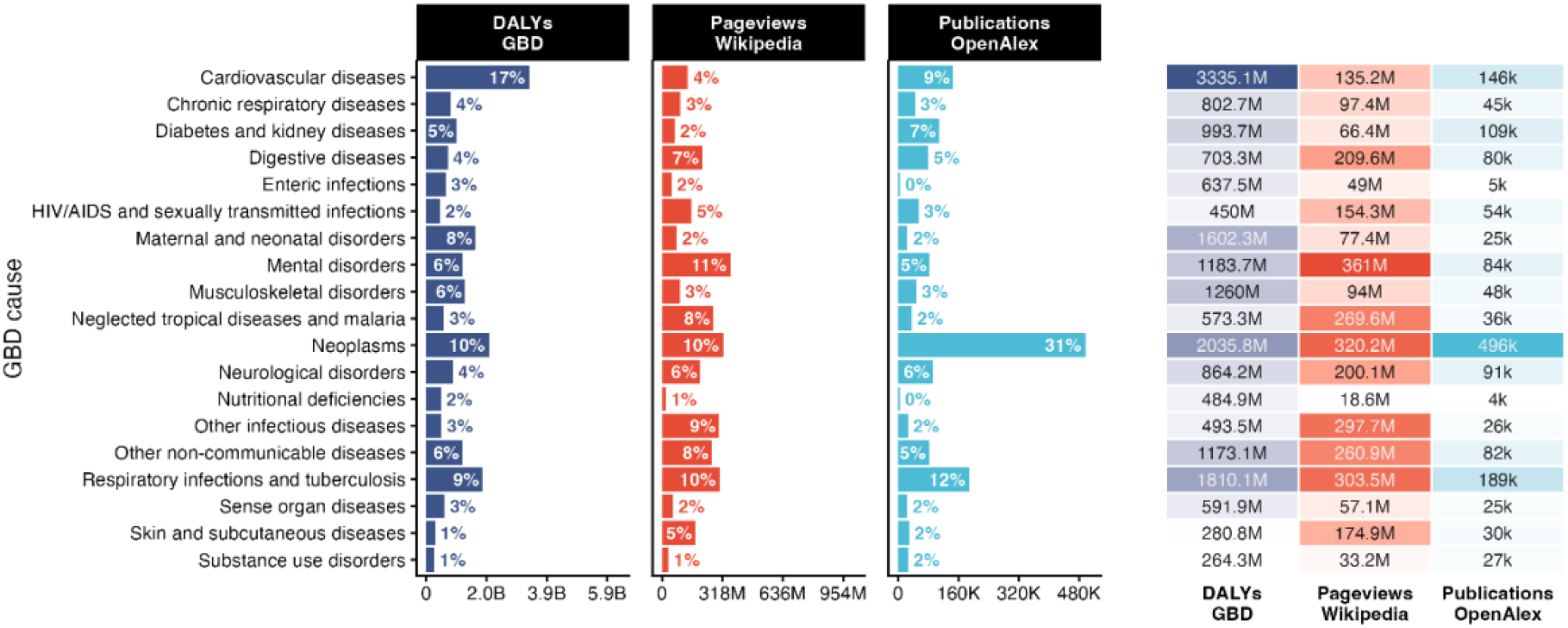
Percentage distribution (left) and the total volume (right) of disease burden, public attention and research efforts across the 19 level-2 GBD disease groups. *Note*: The total volume is the sum of over the study period (2016-2023)

The ratio between the percentage of publications over the percentage of DALYs tells whether research efforts are above (>1) or below (<1) what would be expected based on health needs captured by global disease burden. A ratio above 1 (below 1), means that the disease is over studied (under studied) in relation to its burden. The ratio between the percentage of publications over the percentage of pageviews estimates relative research efforts compared to public attention. By taking both ratios into consideration, one can spot which disease groups are over- or under studied with respect to each of these dimensions of societal demand. For example, cardiovascular diseases are under studied from the perspective of disease burden (ratio = 9.1% / 17.1% = 0.53), whereas they are over studied from the viewpoint of public attention (ratio = 9.1% / 4.3% = 2.14).

The divergences across dimensions become more apparent at the level of individual diseases (level 3) – Fig. S1-S19. Among the 12 level-3 *cardiovascular diseases* (Fig. S1), *ischaemic heart disease* and *stroke* together account for 80% of the DALYs of this category. However, *stroke* is more evenly distributed across the three dimensions, whereas *ischaemic heart disease* concentrates 44% of the burden but only 9% of Wikipedia pageviews and 14% of research publications. Among the 10 *mental disorders* (Fig. S8), *depressive* and *anxiety disorders* concentrate 61% of the DALYs yet attract only 16% of public attention, while *autism spectrum disorders* and *schizophrenia* gather high public attention and concentrate significant research efforts (between 19% and 21%) despite their lower burden (6% and 11%, respectively). Among the 20 *neglected tropical diseases and malaria* (Fig. S10), *malaria* accounts for 72% of DALYs, whereas number of publications and pageviews are distributed more evenly across the category. Within the 34 level-3 *neoplasms* (Fig. S11), research efforts concentrate on *breast cancer* (13%) and *lung cancer* (11%), although the former accounts for only about half the DALYs of the latter. By contrast, *leukaemia* accounts for 17% of the category’s pageviews with a far smaller share of burden and research.

### 3.2. Public attention and disease burden are weakly correlated

Having established that the three dimensions display both alignments and divergences in their distributions, we now examine the statistical relationships between them. Given that public attention and disease burden are two indicators of health demands, if they were highly correlated, public attention would add little additional insight. If instead, the relationship between these variables is weak or mild, then Wikipedia pageviews provide a complementary dimension to understand health demands, which may be helpful for thinking about prioritizing health efforts.

First, we analyse the correlations between dimensions at both hierarchical levels (Figures 3a and 3b). At level-2, based on 19 disease categories, DALYs and pageviews show the weakest correlation (τ = 0.25), while pageviews and publications (τ = 0.46) and DALYs and publications (τ = 0.49) show moderate correlations. At Level 3, with 138 diseases, the correlation between DALYs and pageviews increases slightly (τ = 0.27) and remains weak, while the correlation between pageviews and publications declines clearly (τ = 0.35) and, to a lesser extent, between DALYs and publications (τ = 0.44). These results suggest that public attention, captured through Wikipedia pageviews, captures a different dimension of health demand and that it only partially overlaps with epidemiological burden.

**Fig. 3.**
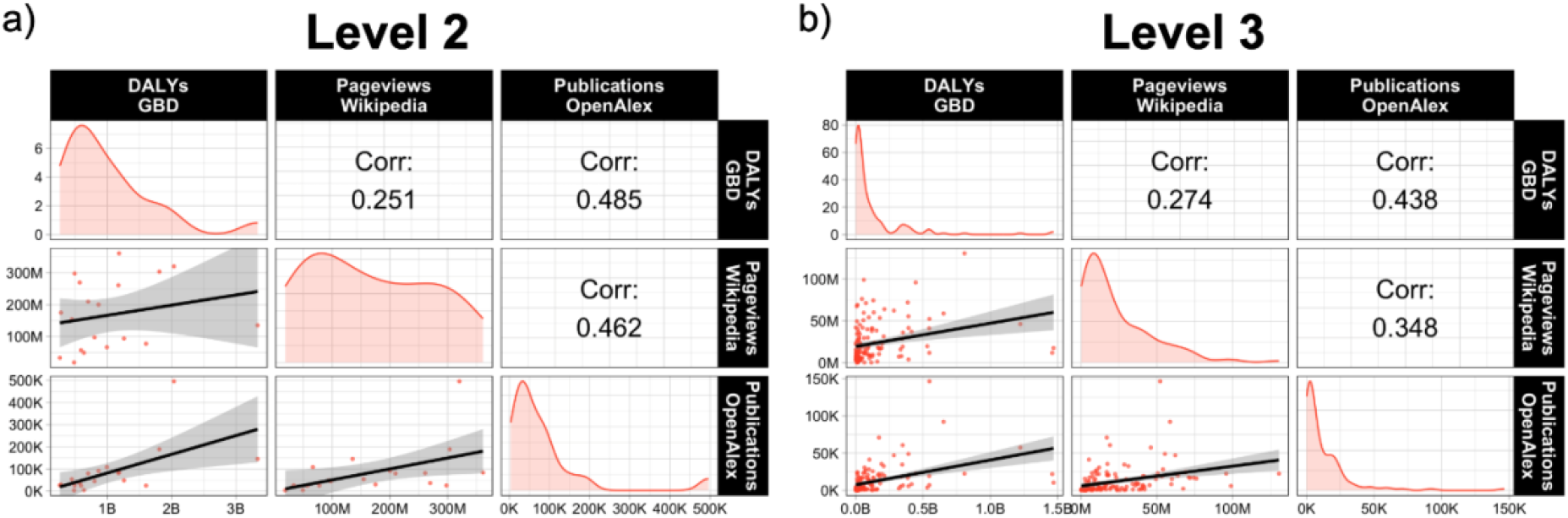
Pairwise correlations between DALYs, pageviews, and publications at disease group and specific disease levels

Another way of exploring whether disease burden and social dimension represent different types of demand is to plot the ratio of science supply over health demands for each of them (see Fig. 4, using level 3). We observe a large dispersion. Some diseases (like most *cancers*) have a relative high ratio in relation both to attention and burden – we can think of this as relative research over supply. Several *infectious diseases* (like *meningitis*, *scabies*, and *measles* affecting mainly Global South countries) have relative low ratios on both sides – this signal a relative research under supply. Yet some diseases (like *rheumatic heart disease, stroke* or *neonatal disorders*) have a low ratio for burden, but a high ratio for public attention (top left of Fig. 4), meaning that there is under supply in relation to burden. And some diseases (like *rabies*, *leishmaniosis* and *inflammatory bowel disease*) have low ratio for attention, but high ratio for burden, signalling under supply in relation to attention.

**Fig 4.**
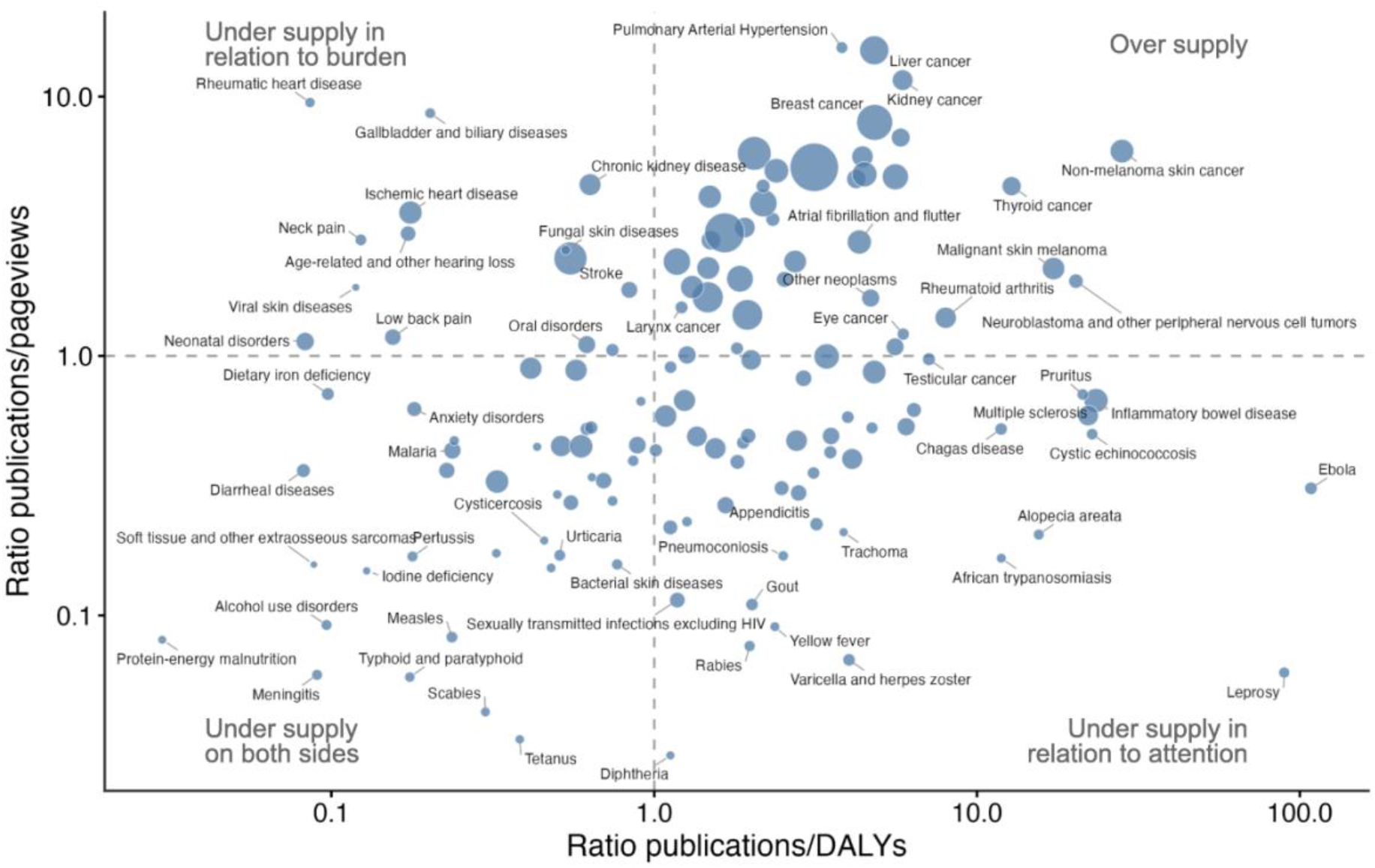
Scatterplot of knowledge supply-to-demand ratios at the level of specific disease. The x-axis shows the ratio as research effort relative to disease burden and the y-axis shows the ratio as research effort relative to public attention

### 3.3. Ternary plots for mapping diseases profiles

Ternary plots are presented at GBD levels-2 and-3 to provide detailed and complementary evidence on the relative patterns of epidemiological burden, public attention to disease and research effort. Fig. 5a shows the ternary plots for the 19 diseases at level-2. Let us remind the reader that the vertical axis signals more (in blue) or less research effort, whereas the horizontal axis shows more health demands in terms of diseases burden (right, in red) or in terms of public attention (left, in green). Some diseases (12) cluster near the central area of the triangle with relatively balanced profiles. However, closer to the vertices, we observe the cases with low balance. At the bottom and the right-hand side, in red, three diseases show a high burden but low research effort and public attention, indicating a DALYs dominant profile, with *nutritional deficiencies* as the most extreme case. At the bottom left, *neglected tropical diseases and malaria*, *skin and subcutaneous diseases*, and *other infectious diseases* are characterized by significant public attention but relatively low research efforts and burden. Going towards the top vertex (publications), as already seen in Fig. 2, *neoplasms* is the disease group where research effort is more dominant.

**Fig. 5.**
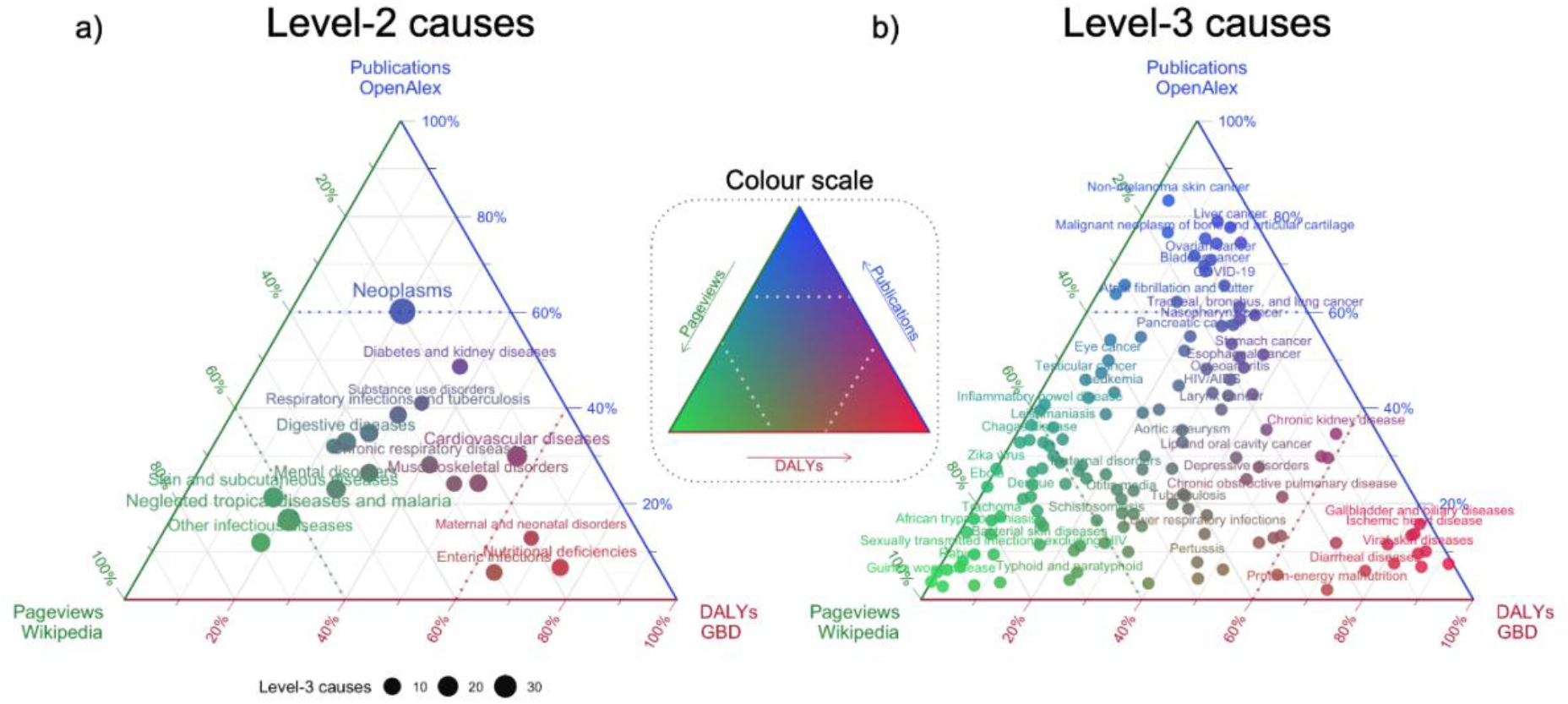
Ternary plot of disease across disease burden, research effort and public attention dimensions. *Note*: The left panel displays Level-2 diseases, where point’s size corresponds to the number of level-3 diseases within each category. The right panel disaggregates these into individual level-3 diseases

In turn, Fig. 5b shows diseases profiles based on GBD level-3. This more granular level, with 138 diseases, reveals a wider variety of patterns, accounting for misalignments across specific diseases. At the bottom right (in red), *rheumatic heart disease* (*cardiovascular diseases*, level-2) and *neonatal disorders* (*maternal and neonatal disorders*, level-2) are characterized by high DALYs but low research efforts and public attention. At the top vertex (in blue), the analysis reveals *liver cancer*, *non-melanoma skin cancer*, and *malignant neoplasm of bone and articular cartilage* as the diseases with the highest research effort in comparison to health needs across the entire level-3. At the bottom left (in green), the disaggregated analysis of *neglected tropical diseases and malaria* shows that *African trypanosomiasis*, *Chagas disease*, *rabies*, and *trachoma* are extremely skewed towards the public attention vertex, signalling that public attention far exceeds the research efforts devoted to them. At the bottom in the centre in Fig. 5b, there are those conditions that present high burden and public attention but low research effort such as *iodine deficiency, lower respiratory infections*, and *meningitis*, among others. Finally, diseases near the central region of the triangle are distributed more evenly across three dimensions, representing the closest approximation to balanced alignment. At the level of disease groups (level-2), this zone is more densely populated, with *respiratory infections and tuberculosis* being the most balanced (Fig. 5a). However, at level-3 disaggregation, few diseases (e.g. *aortic aneurysm*) show balance between burden, research effort and public attention.

Fig. 6 details diseases at level-3 for four broad disease categories with positions relative to those in their disease group at level-2 and relative to all diseases. The arrows indicate the extent to which the specific pattern of each level-2 differs from the overall pattern across all diseases. This visualisation makes it easier to identify misalignments and outliers within each disease group. Within *cardiovascular diseases*, *hypertensive heart disease*, and *atrial fibrillation and flutter* receive comparatively greater attention than would be expected given their corresponding research efforts and burden. *Stroke*, by contrast, shows a more balanced profile. Within *mental disorders*, patterns shift slightly towards the research effort vertex, but no single disease stands out in terms of the number of publications relative to its burden and public attention. Similarly, *neglected tropical diseases* show a slightly more balanced profile. However, this more granular comparison reveals important differences: *Chagas disease* and *leishmaniosis, are* over supply compared with other neglected diseases, while *malaria* is clearly under supply in relation to its high disease burden. The analysis for *neoplasms* reveals that all cancers stand out in terms of research efforts when compared with other all diseases (Fig. 6), but the group disease analyses reveals that several are undersupplied in relation to public attention such as *mesothelioma*, *testicular cancer* and *malignant skin melanoma*.

**Fig. 6.**
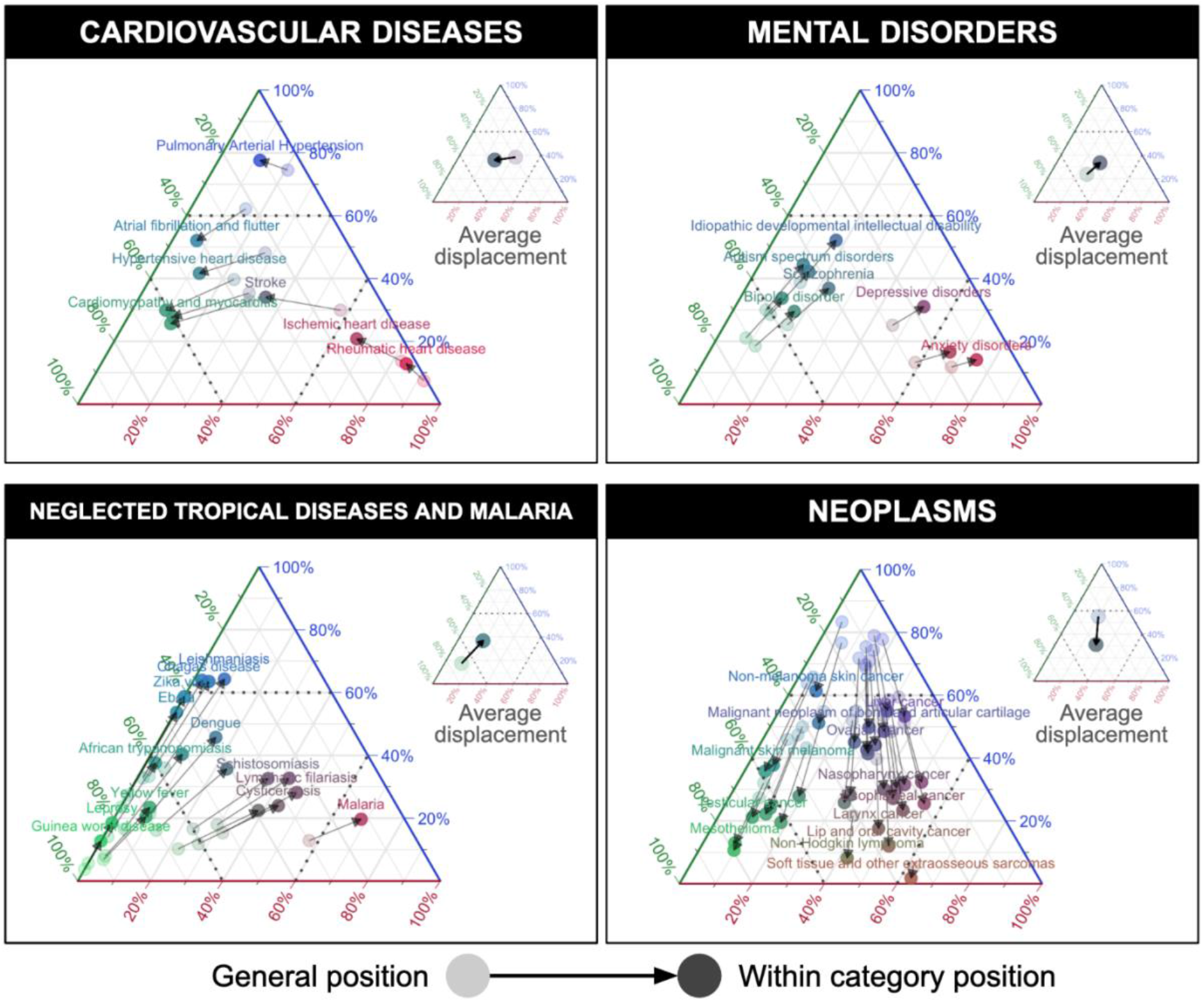
Ternary plots of specific diseases with normalisations within the disease group (near arrow tip) and for all diseases (faded)

### 3.4. Territorial disaggregation

While the previous results capture global world patterns, we conduct the analysis on a finer territorial scale to account for the particularities of different geographical contexts. As Wikipedia is organized around language editions rather than countries, we have selected four linguistic areas that reflect specific territories in the Global North and Global South and for which social and cultural differences are expected: German (133 million speakers), Persian (127 million speakers), Swahili (90 million), and Vietnamese (97 million).

Fig. 7 presents the burden, public attention, and research effort profile of each region, alongside ternary plots at level-2. *Cardiovascular diseases* lead the burden dimension in the German, Persian, and Vietnamese linguistic areas (ranging from 16% to 21%), consistent with the global pattern. However, this share is considerably lower for Swahili speakers (6%). *Neoplasms* concentrate the greatest research efforts in the German, Persian, and Vietnamese areas, though the number of publications ranges considerably from 22% to 34%. In contrast, research efforts in the Swahili speaking area concentrate on *HIV/AIDS and sexually transmitted infections* (32%) which, in turn, are also the diseases that gather the most public attention (19%).

**Fig 7.**
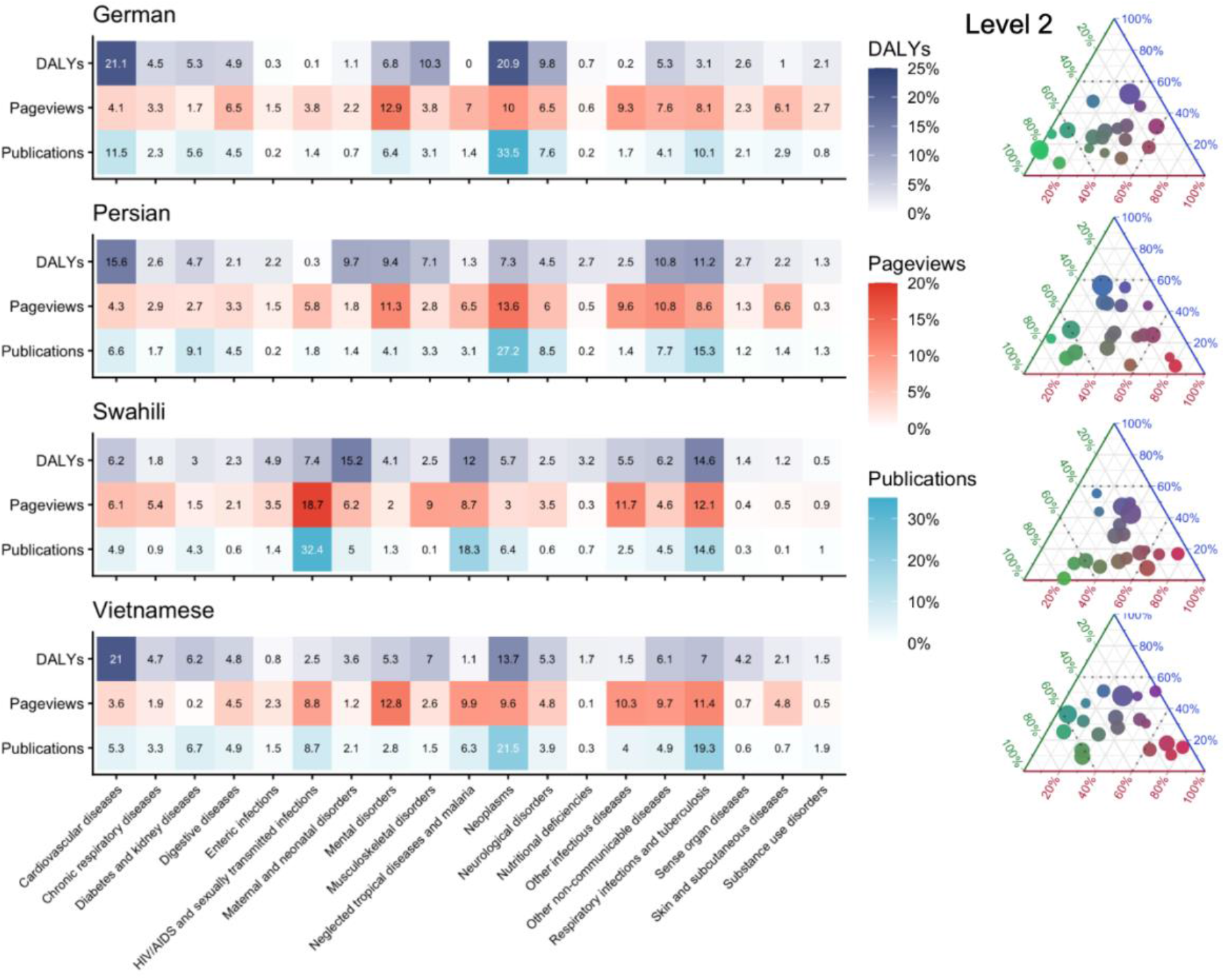
Territorial distribution of disease groups (level-2) across four linguistic areas. *Note*: The left panel displays shares of DALYs, pageviews, and publications for disease groups. The right panel presents the ternary plots

Overall, public attention is heavily concentrated in the Swahili speaking area, with three diseases each receiving more than 10% of attention and together accounting for 42% of the total number of Wikipedia page views. A similar pattern can be found in the Vietnamese speaking area. In contrast, public attention is more scattered in the German speaking area with only two diseases groups reaching 10% of public attention to disease. It is worth noting that *mental disorders* account for a significant share of Wikipedia pageviews in the Vietnamese (13%), German (13%), and Persian (11%) language areas, reflecting social aspects of disease not captured by DALYs. A similar pattern applies to *skin and subcutaneous diseases*.

Looking at more area specific patterns, *respiratory infections and tuberculosis* shows different profiles across regions. In the Vietnamese speaking area, this category represents a moderate share of burden (7%) but rises to second place in both public attention (11%) and research efforts (19% of the total number of publications). In the Persian speaking context, by contrast, it ranks second in burden (11%) and research efforts (15%), but occupies a less prominent position in public attention (9%), whereas in the Swahili speaking area it is prominent across the three dimensions, ranking second in burden (14%) and pageviews (12%), and third in research effort (15%).

Finally, Fig. 8 illustrates territorial patterns for four selected diseases that show distinct supply-demand profiles across the German, Persian, Swahili, and Vietnamese linguistic areas. Each disease is shown in a ternary plot, with the global pattern represented by a large circle and the four linguistic areas by smaller labelled points. This reveals interesting local imbalances that remained invisible. In the case of *stroke*, which its overall patterns shows under supply in relation to burden (see Fig. 4), all four regions cluster close to the global pattern, with some small divergences, particularly in research effort. In the Vietnamese speaking area, the burden is higher (75%) and the research effort lower (10%) with regard to the overall pattern (burden: ≈60%, research effort: 30%), whereas public attention is similar across the four linguistic areas (10-20%). Greater research effort is devoted in the German area (40%) despite lower burden (50%). By contrast, *liver cancer,* which is globally over supplied, shows a Global North-South divide. The German speaking area follows the general pattern (research effort: 75%, burden: 20%, public attention: 5%), whereas the Persian speaking area concentrates lower research effort (60%), despite similar burden (20%) and higher public attention (20%). The Swahili case is more acute, with negligible research efforts (10%) despite considerable social demand (50%) and twice the burden (40%). In the case of *alcohol use disorders*, under supply in relation to burden and attention, research effort is consistently low (10%) across the four linguistic areas. Notably, the German and Vietnamese linguistic areas show higher burden (60% compared with the global pattern of ≈50%) but lower social attention. Finally, the territorial patterns for *rabies*, whose global pattern is characterized by undersupply in relation to public attention, reveal extreme imbalances across territories. The German and Persian language areas follow the global pattern, with very high public attention (≈90%) and negligible research effort. This contrasts with the pattern observed for the Vietnamese and Swahili-speaking areas. In the former case, there is an oversupply of publications despite low public demand and low health demand. In the latter case, public attention (50%) is relatively matched by research effort (40%). The lower disease burden (10%) may reflect common concerns about the fragmented and incomplete GBD coverage of these conditions.

**Fig 8.**
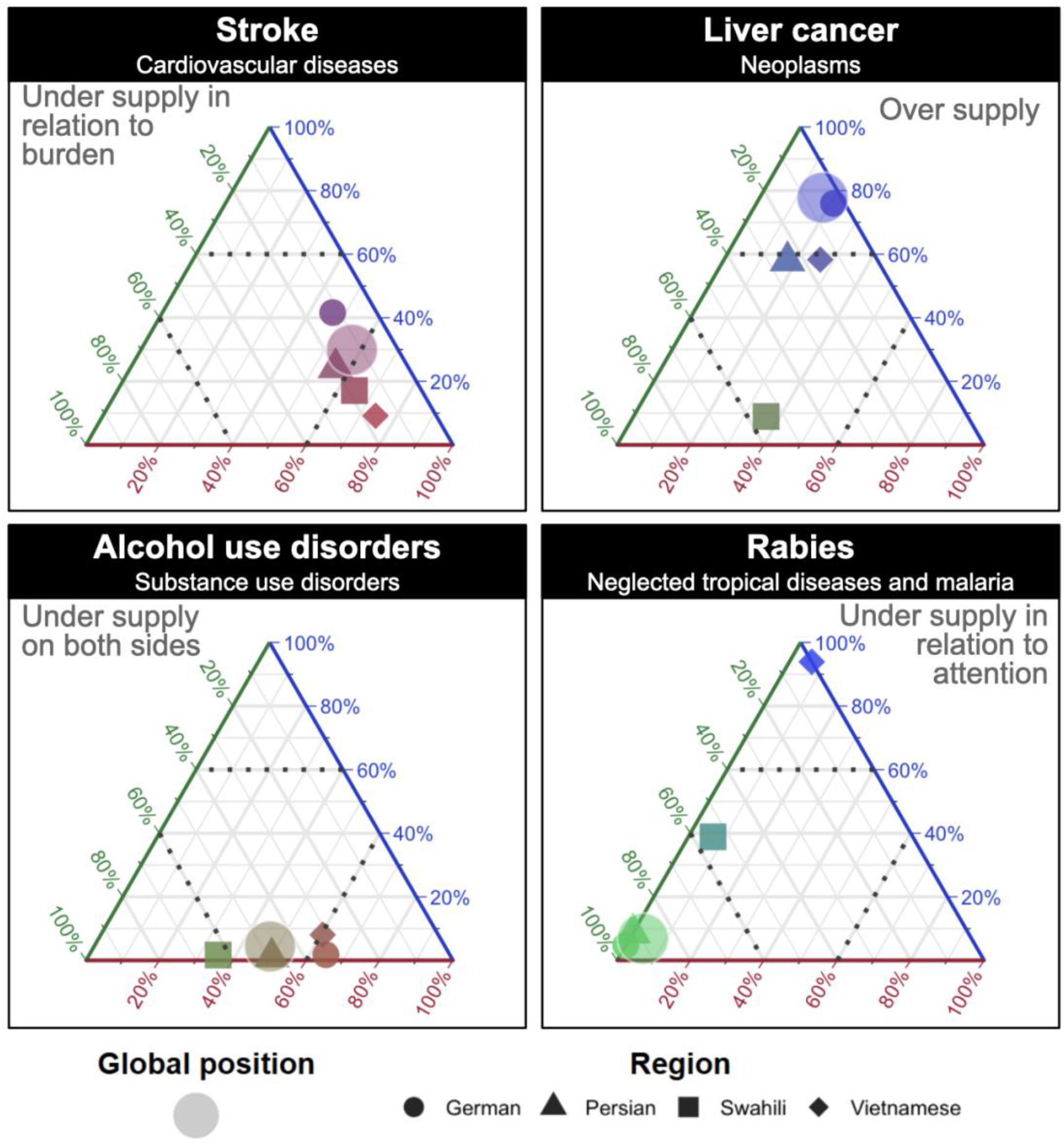
Ternary plots of four specific disease (level-3) across four selected linguistic areas, compared against the global aggregate.

## 4. Discussion and conclusions

Our findings show limited alignment between the supply of scientific knowledge and health needs and demands, not only in terms of disease burden, but also in terms of public attention. The study also demonstrates that public attention captures a different dimension of health demand than epidemiological burden. Some diseases attract public attention beyond what their DALYs alone would suggest, pointing to forms of concern, visibility, stigma, or information seeking. At the disease group level, cardiovascular diseases account for the largest share of disease burden, mental disorders attract the largest share of public attention, and neoplasms concentrate by far the largest share of research effort. At the level of specific diseases, our analysis reveals substantial variation within disease groups. Territorial analyses further show distinct profiles across language areas and Global North-South differences.

Our study makes two contributions. First, drawing on Sarewitz and Pielke’s [13] supply-demand framing of science policy, we extend existing work on disease burden and research effort by treating public attention as a second form of health demand. This allows us to compare knowledge supply, captured by research effort, with both epidemiological demand, captured by disease burden, and social demand, captured by public attention to disease. Second, we use ternary plots as a mapping technique to identify where knowledge supply aligns, or fails to align, with these two forms of demand, and which dimension drives the imbalance. They can be presented at different levels of disease aggregation, each revealing different nuances. To support transparency and reproducibility, we make the disease mapping, data, and code openly available.

This study has several limitations. First, Wikipedia access depends on internet access, language, digital literacy, and the availability and quality of articles, which likely over represent populations in the Global North. Although progress in internet access has been faster in Africa than in any other continent [56], access remains uneven and still lags behind the global average [57]. Second, Wikipedia data are organised by language edition rather than by country. Our territorial results are therefore linguistic rather than national. A single language edition draws readers from several countries, and most countries host more than one language. Third, matching diseases across the GBD classification, MeSH terms, and Wikipedia articles involves judgment because there is no one-to-one correspondence between the sources. Nevertheless, our research design shows near complete overlap with available previous studies [10]. Fourth, research effort is measured through publications, which do not capture all forms of knowledge supply, such as financial amount of funding or medical innovation (e.g. clinical trials). Similarly, public attention is broader than Wikipedia use.

### 4.1. Policy implications and future research

Our approach helps health priority-setting move beyond a simple comparison between research effort and disease burden. By mapping knowledge supply against both epidemiological and social demand, it allows to more fully explore where research effort is misaligned. These different demands and forms of misalignment may raise different health policy questions and require different responses.

Coburn et al. [19] and Kumar et al. [26] discussed that some high-burden conditions (such as maternal health) require local or national improvements in healthcare systems or prevention rather than more research, whereas others are under researched (such as malaria) both locally and globally. Thus, certain imbalances may call for disease-specific approaches such as targeted research, a public health campaign or vaccination (vertical interventions), while others may point instead to strengthening countries’ research capacity and healthcare systems that operate across diseases groups (horizontal interventions). In particular, Coburn et al. [19] make this point in relation to vertical interventions: while disease-specific targeting can mobilize resources, it can also focus attention too narrowly on research and innovation, leaving local research capacity and health system strengthening underdeveloped. Horizontal interventions, however, are harder to monitor and evaluate economically and epidemiologically and tend to have more downstream impact [17,58].

The science supply-health demand framework is not only relevant to academic debates but is also being discussed in research policy practice [59]. Our mapping of supply-demand through ternary plots can be particularly useful to compare diseases across geographical contexts. In the Global North, our framework can help identify whether health demand is driven mainly by public attention, disease burden, or both. These different patterns may point to different responses. When a disease is undersupplied in relation to burden, as in cardiovascular diseases or stroke, responses need to focus on prevention, treatment and healthcare. However, forms of health demand driven by public attention that are not necessarily among countries’ most pressing domestic health needs may require action more oriented toward public awareness and health literacy (e.g. mental disorders in the German speaking area) or remain relevant for international cooperation and development funding (e.g. tuberculosis and malaria).

In the Global South, the approach can help identify diseases or disease groups where high burden is much higher than research effort possibly because knowledge supply already exists (e.g. diarrheal infections) or conditions that require greater efforts despite not having a high burden (e.g. neglected tropical diseases). This makes public attention particularly valuable in Global South contexts for conditions such as neglected tropical diseases, where DALYs may underestimate the full burden because of weaker surveillance systems and limited diagnostic capacity [21,60].

For example, in the Swahili-speaking area, HIV/AIDS and sexually transmitted infections are not under researched: they concentrate the largest share of research effort among all disease groups (Fig. 7). At the same time, they attract substantial public attention and account for a considerable burden, with a DALY share at least three times higher than in the German, Persian, and Vietnamese areas. This pattern occurs despite the significant global response and progress made through vertical interventions, including expanded access to HIV/AIDS care and treatment [58]. Yet the persistence of relatively high burden despite high research effort suggests that horizontal interventions may need to be prioritized to strengthen healthcare systems and local innovation capacity. In fact, increasing research capacity in low-income regions has been proposed as a strategy to address local health needs once effective treatments exist and burden has been reduced but is still sizeable [14].

Unlike HIV/AIDS, neglected tropical diseases and malaria receive much lower research effort, while attracting considerable public attention not only in Global South regions where burden is high but also across other areas, as shown by our territorial analysis (Fig. 7 and 8). This pattern may point to the need for continued vertical interventions, including targeted research. At the same time improving broader living and health conditions, including access to clean water, sanitation, and primary healthcare is crucial for prevention of non-communicable diseases [21,24]. Fuady et al. [61] identify coordination and collaboration across sectors and institutions and flexible use of funds as central gaps for improving broader living conditions.

Future work could extend our framework in two directions. First, it could incorporate other indicators of the social dimensions of disease, such as patient organization activity, media coverage, or advocacy and disease-specific campaigns. Second, it could move from descriptive mapping to explanatory analysis by examining whether epidemiological burden and public attention help explain variation in scientific knowledge production (publications, clinical trials) across diseases. For example, future studies could test whether public attention increases research effort above the effect ‘predicted’ by disease burden, or whether it moderates the relationship between disease burden and publication and medical innovation outputs. For example, low public attention to cardiovascular diseases may help account for its lower research effort despite its high burden, whereas high levels of public attention to skin diseases may help explain their higher levels of research effort. This question may be particularly relevant in high-income contexts, such as the German linguistic area, where public attention could play a stronger role in shaping research priorities, given that it is likely to be associated with market demand.

### 4.2. Concluding remarks

Public attention provides a complementary dimension for mapping global health needs and demands. By combining disease burden, research effort, and public attention, this study identifies where knowledge supply fails to match epidemiological and social demand, and which dimension drives the imbalance. Our framework supports more nuanced global health priority-setting by making visible the different forms of (mis)alignment of knowledge supply with demands from both epidemiological burden and societal needs.

## Ethics approval and consent to participate

Not applicable.

## Consent for publication

Not applicable.

## Availability of data and materials

The complete disease mapping is publicly accessible via Zenodo (https://doi.org/10.5281/zenodo.21694438), and all processing scripts, code, and documentation are available on GitHub (https://github.com/Wences91/gbd_misalignment). Supplementary material is available at: https://doi.org/10.5281/zenodo.21709258.

## Authors’ contributions

ADF conceived the original idea. All authors contributed to the framing of the study and participated in the literature review. WAM constructed the dataset by integrating the three data sources and performed the analyses and visualizations. All authors contributed to drafting the manuscript. ADF and IR revised the manuscript substantially. All authors read and approved the final manuscript.

## Acknowledgements

We thank Alysson Mazoni and the University of Campinas for providing access to the OpenAlex publications in-house database. We are grateful to previous researchers on this topic, in particular Alfredo Yegros [9, 26], for making classifications of their articles fully transparent and accessible.

## Footnotes

1 GBD is noted for having some limitations in terms of data quality: accuracy, completeness, the diversity of data sources and how data are processed [5].

2 https://en.wikipedia.org/wiki/Censorship_of_Wikipedia

3 https://en.wikipedia.org/wiki/Malaria

